# An in-depth statistical analysis of the COVID-19 pandemic’s initial spread in the WHO African region

**DOI:** 10.1101/2021.08.21.21262401

**Authors:** Ananthu James, Jyoti Dalal, Timokleia Kousi, Daniela Vivacqua, Daniel Cardoso Portela Câmara, Izabel Cristina dos Reis, Sara Botero-Mesa, Wingston Ng’ambi, Papy Ansobi, Beat Stoll, Cleophas Chimbetete, Franck Mboussou, Benido Impouma, Cristina Barroso Hofer, Flávio Codeço Coelho, Olivia Keiser, Jessica L Abbate

**Affiliations:** Department of Chemical Engineering, Indian Institute of Science, Bangalore, India; Institute of Global Health, Faculty of Medicine, University of Geneva, Geneva, Switzerland; Department of Pediatric Infectious Diseases, UFRJ, Rio de Janeiro, Brazil; Laboratório de Mosquitos Transmissores de Hematozoários - LATHEMA, Instituto Oswaldo Cruz, Fundação Oswaldo Cruz, Rio de Janeiro, Brazil; Núcleo Operacional Sentinela de Mosquitos Vetores - NOSMOVE, Fundação Oswaldo Cruz, Rio de Janeiro, Brazil; Health Economics Policy Unit, Department of Health Systems and Policy, College of Medicine, University of Malawi, Lilongwe, Malawi; Research and Training Unit in Ecology and Control of Infectious Diseases (URF-ECMI), Faculty of Medicine, University of Kinshasa, Kinshasa, Republic Democratic of Congo; Newlands Clinic, Harare, Zimbabwe; World Health Organization, Regional Office for Africa, Brazzaville, Congo; School of Applied Mathematics, Getulio Vargas Foundation, Rio de Janeiro, Brazil; UMI TransVIHMI (Institut de Recherche pour le Développement, Institut National de la Santé et de la Recherche Médicale, Université de Montpellier), Montpellier, France; Geomatys, Montpellier, France; The Global Research and Analysis for Public Health (GRAPH) Network (https://thegraphnetwork.org), Association Actions en Santé, Geneve, Switzerland

**Author notes:** **Corresponding author:** Ananthu James. Shared first authorship (Ananthu James, Jyoti Dalal, Timokleia Kousi). Shared senior authorship (Olivia Keiser, Jessica L Abbate).

**Keywords:** COVID-19, Epidemiology, Public health, sub-Saharan Africa, SARS-CoV-2, Emerging Infectious Disease, Comparative analysis, Cross-sectional study

## Abstract

During the first wave of the COVID-19 pandemic, sub-Saharan African countries experienced comparatively lower rates of SARS-CoV-2 infections and related deaths than in other parts of the world, the reasons for which remain unclear. Yet, there was also considerable variation between countries. Here, we explored potential drivers of this variation among 46 of the 47 World Health Organization African region member states in a cross-sectional study. We described five indicators of early COVID-19 spread and severity for each country as of 29 November 2020: delay in detection of the first case, length of the early epidemic growth period, cumulative and peak attack rates, and crude case fatality ratio (CFR). We tested the influence of 13 pre-pandemic and pandemic response predictor variables on the country-level variation in the spread and severity indicators using multivariate statistics and regression analysis. We found that wealthier African countries, with larger tourism industries and older populations, had higher peak (p < 0.001) and cumulative (p < 0.001) attack rates, and lower CFRs (p = 0.021). More urbanized countries also had higher attack rates (p < 0.001 for both indicators). Countries applying more stringent early control policies experienced greater delay in detection of the first case (p < 0.001), but the initial propagation of the virus was slower in relatively wealthy, touristic African countries (p = 0.023). Careful and early implementation of strict government policies were likely pivotal to delaying the initial phase of the pandemic, but did not have much impact on other indicators of spread and severity. An over-reliance on disruptive containment measures in more resource-limited contexts is neither effective nor sustainable. We thus urge decision-makers to prioritize the reduction of resource-based health disparities, and surveillance and response capacities in particular, to ensure global resilience against future threats to public health and economic stability.

**Summary Box:** *What is already known on this topic?:*

- COVID-19 trajectories varied widely across the world, and within the African continent.
- There is significant heterogeneity in the surveillance and response capacities among WHO African region member states.

*What are the new findings?:*

- Cumulative and peak attack rates during the first wave of COVID-19 were higher in WHO African region member states with higher per-capita GDP, larger tourism industries, older and more urbanized populations, and higher pandemic preparedness scores.
- Although better-resourced African countries documented higher attack rates, they succeeded in limiting rapid early spread and mortalities due to COVID-19 infection.
- African countries that had more stringent early COVID-19 response policies managed to delay the onset of the outbreak at the national level. However, this phenomenon is partially explained by a lack of detection capacity, captured in low pandemic preparedness scores, and subsequent initial epidemic growth rates were slower in relatively well-resourced countries.

*What do the new findings imply?:* Careful implementation of strict government policies can aid in delaying an epidemic, but investments in public health infrastructure and pandemic preparedness are needed to better mitigate its impact on the population as a whole.

## Introduction

The first confirmation of a COVID-19 case in the African continent occurred in Egypt on 14 Feb 2020 [1]. Following that introduction, along with others that would go on to occur one-by-one in the rest of the continent, COVID-19 cases and related fatalities rose exponentially, eventually reaching all African countries. Countries on the continent appear to have fared better during the initial wave of the pandemic than elsewhere in the world, with lower attack rates and many orders of magnitude fewer deaths. There are various theories about the drivers of this phenomenon, including younger population structure, lower rates of obesity and other comorbidities, higher rates of immune-modulating parasitic diseases, relatively low population densities and urbanization, and lower pandemic preparedness and detection capacities in resource-limited contexts [2-6]. While the as-yet-undetermined cause of this difference in confirmed COVID-19 cases and deaths setting African countries apart could be shared across the continent, there was nevertheless heterogeneity in the evolution of the epidemic between countries [7]. It is important to understand the factors that may contribute to the differences among these countries, as these insights can inform policy and priorities for future public health emergencies when comparison to the rest of the world is marred by problematic biases [8].

Motivated by this objective to examine drivers of heterogeneous COVID-19 spread and severity across African countries, we quantified how the first wave of the COVID-19 pandemic unfolded in the 47 member states comprising the World Health Organization (WHO) African region using data extracted from official country health ministry announcements and published daily on the WHO COVID-19 dashboard [9]. We then performed a statistical analysis of the relationship between indicators of COVID-19 spread and severity and pre-pandemic socioeconomic and demographic aspects specific to each country, as well public health policy characteristics both prior to and during the pandemic. We end the study with an interpretation of these relationships, and suggestions for how the results might be used to help decision-makers improve future epidemic preparedness and response.

## Methods

### Study design and settings

This is a cross-sectional analysis of the COVID-19 data reported from the start of the pandemic through 29 Nov 2020 (inclusive) among the 47 member states comprising the WHO African region. We analyzed the data gathered by the WHO [9] on the daily number of new cases and deaths published by country health ministries through official channels. This period captures the initial wave of the pandemic for most countries in the region, ending prior to complications driven by the festive season and roll-out of vaccines. While members of our team were responsible for compiling these data with the WHO during this time, all data were systematically published to the WHO COVID-19 dashboard [9] and are thus freely accessible. We note that groups of confirmed cases marked as ‘probable’ due to sole availability of rapid diagnostic (viral antigen) tests - representing 70 cases from Comoros and one case from the Democratic Republic of Congo (DRC) - are now also considered part of the official case counts and were included as such in the present study. We included the cases with information about patient outcome status (alive, recovered, or dead), excluding those with missing values (8 from Burundi, 4 from Comoros, and 3 from Rwanda). Because data from the Republic of Tanzania were not shared during this period, our analysis focused on the remaining 46 countries (Figure 1).

**Figure 1.**
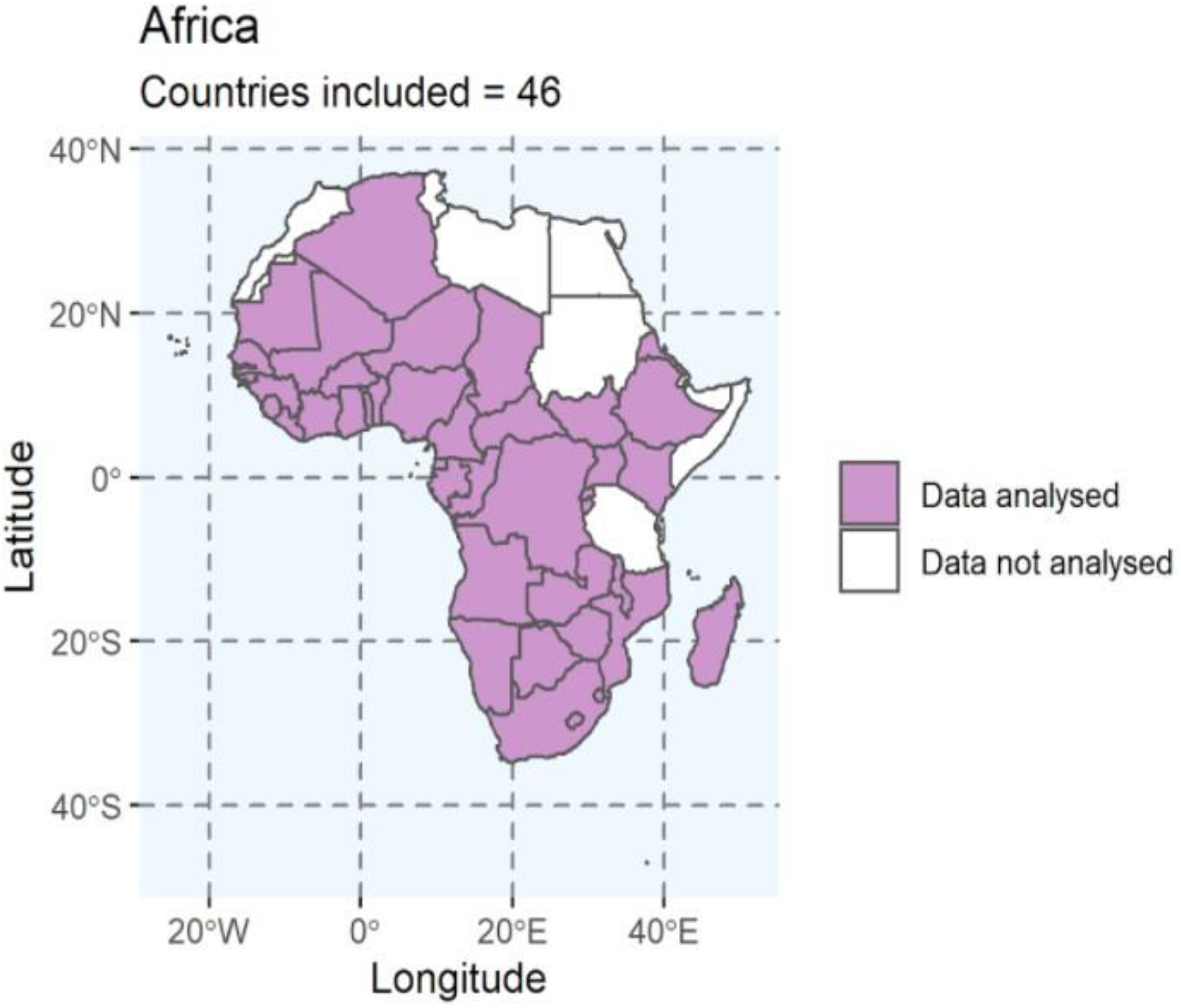
The 46 countries from the WHO African region included in this study. We excluded The Republic of Tanzania because of underreporting as the last case reported publicly was on 7 May 2020.

Predictor variables were pulled from publically available datasets (WHO, World Bank, IndexMundi, etc.), for which the complete set of specific sources is provided in Supplementary Information Table S1. Data were processed using both Python (version 3.7.10) and R (version 4.0.2) (https://www.r-project.org/). All statistical analyses were performed in R (version 4.0.2).

### Patient and public involvement

Neither patients nor the public were involved in the design, conduct, reporting, or dissemination plans of our research.

### Data

#### Epidemic spread and severity indicators

We included five response indicators describing the size, severity, and evolution of the outbreak both within each country and in the context of the whole WHO African region. For each of the 46 countries, these included: 1) the cumulative attack rate (cumulative number of cases per million inhabitants); 2) the maximum monthly attack rate (new cases per million inhabitants summed over consecutive 4-week intervals); 3) the crude case fatality ratio (CFR, the ratio of the cumulative number of deaths to the cumulative number of cases, not accounting for lags in death or reporting of deaths); 4) the relative delay in which the epidemic reached each country, the *start delay*, expressed as the number of days between the first reported case in the country and the first reported case in the region (on 25 February 2020 in Algeria); and 5) the length of the early epidemic growth phase, the *initial growth period*, measured as the number of days between the date of reporting of the first and the 50th case in the country (also expressed as the inverse of the *initial epidemic growth rate* in the country).

The relative epidemic start delay was motivated by Li et al., 2021 [10], representing a delay in either the exposure, testing capacity, data reporting, or some combination thereof. The length of the initial epidemic growth period, the inverse of the initial epidemic growth rate, was *a priori* expected to correlate with population density given that respiratory pathogens like SARS-CoV-2 follow density-dependent transmission patterns [11]. We did not attempt to estimate the classical reproduction number, as we were unable to distinguish between new imported cases versus new cases due to community transmission.

#### Predictor variables

To explain the variation in the response variables between countries, we collected 13 pre-pandemic and pandemic response predictor variables from public data repositories (described in detail in Table S1 of the Supplementary Information). These included: per capita Gross Domestic Product (GDP), size of the fishing industry (per capita fishing volume), proportion of the population under 15 years of age, proportion of the population that is male, population density, proportion of population living in urban areas (urbanization), latitude, combined cumulative attack rate in neighboring countries, per capita revenue generated from tourism, per capita number of tourist arrivals, COVID-19 government response stringency index score at the time of reporting of the first case [12], mean stringency index score calculated as the difference between the mean and minimum stringency index scores over the course of the study period, and the country’s self-assessed level of epidemic and pandemic preparedness [13]. We included the predictors on tourism, agriculture, and fishing, as these are the main industries in many of the sub-Saharan African nations (for instance, the island nations [14]). Moreover, the international movement of people for tourism or work-related activities has been documented as an important factor in the spread of the virus at the beginning of the epidemic [15,16]. We used the most recent values for each predictor for the latest period prior to the start of the pandemic (available as of November 2020) [17,18].

### Statistical Analysis

#### Missing data and imputation

We identified 11 missing values in our dataset corresponding to (i) initial and mean government response stringency index for Comoros, Equatorial Guinea, Guinea-Bissau and Sao Tome & Principe and (ii) per capita tourist arrivals from Equatorial Guinea, Liberia, and South Sudan. To preserve inclusion of these countries in our analysis, we used data imputation methods to artificially complete the missing records by substituting the missing values corresponding to the explanatory variables. Per capita tourist arrivals and mean government response stringencies were imputed using the mean values. For initial government response stringency, we substituted the missing values with the value computed using Equation 1:

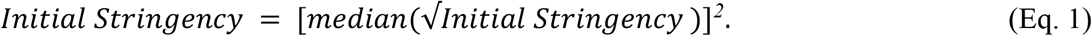

For the cumulative attack rate in neighboring countries for which data were not available via the WHO database, we sourced this information from the freely available Our World In Data (OWID) COVID-19 database for the total number of positive cases (https://ourworldindata.org/covid-cases) for the dates corresponding to the study period.

#### Collinearity and Transformations

Following data imputation, we explored the collinearity of response indicators and predictor variables using parametric Pearson’s correlation matrices. In order to meet the normality assumption of this parametric statistical test, we log-transformed (to base 10) skewed predictor variables (for the per capita number of tourist arrivals, per capita tourism revenue, attack rate of neighboring countries, per capita GDP, per capita fishing volume, and population density). We used a square-root transformation for the initial government response stringency, as it approximated adequately thanks to smaller amplitude in the variance. For the response variables, we (base-10) log-transformed the cumulative attack rate, maximum monthly attack rate, and the initial epidemic growth period; square-root transformed the relative start delay; and performed arcsine square-root transformation on CFRs, as this transformation is most appropriate for proportions bounded by zero and 1 [19].

#### Principal Component Analysis (PCA)

We performed a principal component analysis (PCA) on the full set of untransformed predictor variables to reduce the number and collinearity of variables for statistical analysis. This method performs a linear transformation on the variables and reduces variance in collinear variables to a set of orthogonal components, to which each receives some contribution from the underlying variables. These “composite” variables, or principal component dimensions (PC1, PC2, etc.), can then be used for statistical analysis against response variables. We scaled variables to unit variance using the “PCA” function from the *FactoMineR* package [20]. The resulting informative principal component (PC) dimensions were selected following the Kaiser criterion (eigenvalue > 1) [21]. PCA loadings were used to verify the direction and contribution of each variable when interpreting the resulting dimensions. The scores of the resulting informative dimensions were used as explanatory variables in the regression analysis.

#### Regression analysis

We performed regression analyses to understand the impact of the composite predictor variables (informative PCA dimensions with eigenvalue > 1) derived above on each of the epidemic spread and severity (response) indicators. We modeled each response indicator as count data offset by respective denominators (population size for attack rates, and total cases for death counts to model CFR) in a generalized linear model (GLM) with a negative binomial error distribution using the “glm.nb” function (*MASS* package) following inspection of model diagnostics with alternative error distributions specified. The residual distributions for the best fitting model are provided in Supplementary Information Figure S1. We performed model selection and significance testing using the “stepAIC” function from the *MASS* package, which selects the best model based on the Akaike Information Criterion and measures significance of predictor variables in the best-fit model against the z-test statistic. Statistical significance was considered at p < 0.05. The false discovery rate due to multiple tests, calculated using the Benjamini-Hochberg method (“p.adjust” function), was considered for interpretation of results.

#### Robustness to variation in data quality

In order to check the robustness of our results to data quality issues, we assumed countries with fewer than 10 reported deaths as of 29 Nov 2020 to be suspected of under-detection or under-reporting of cases and deaths. Thus, we repeated the above analyses after removing the four countries that had reported fewer than 10 deaths (i.e., Burundi (deaths = 1), Comoros (deaths = 7), Eritrea (deaths = 0), and Seychelles (deaths = 0)) to check for qualitative differences in the results. As with the full dataset, the negative binomial model was the most appropriate model for each of the response variables.

## Results

### Epidemic spread and severity indicators

The COVID-19 response indicators showed substantial variation across the 46 WHO African region member states included in the analysis (Figure 2A; Figure S2A). The cumulative attack rate ranged from 58.97 in Burundi to 19140.62 in Cabo Verde (median: 886.67). The maximum monthly attack rate varied from 14.83 in Burundi to 4843.33 in South Africa (median: 247.48). Case fatality ratio ranged from 0.00% in Seychelles and Eritrea to 6.00% in Chad (median: 1.76%). The delay in detecting the first case in each country relative to the region (*start delay*),ranged from 0 in Algeria to 74 days in Lesotho (median: 20 days). Finally, the length of the initial epidemic growth period (inverse of the *initial epidemic growth rate*)varied from 8 days (or 6.1 cases per day) in Mauritius to 105 days (equivalent to 0.47 cases per day) in the Gambia (median: 26.5 days). The cumulative and maximum attack rates were highly positively correlated (*r* = 0.9885, p < 0.001; Figure S3). Additional trends for correlation between indicators were observed, notably negative correlations between CFR and attack rates, but none were statistically significant (Figure S3).

**Figure 2.**
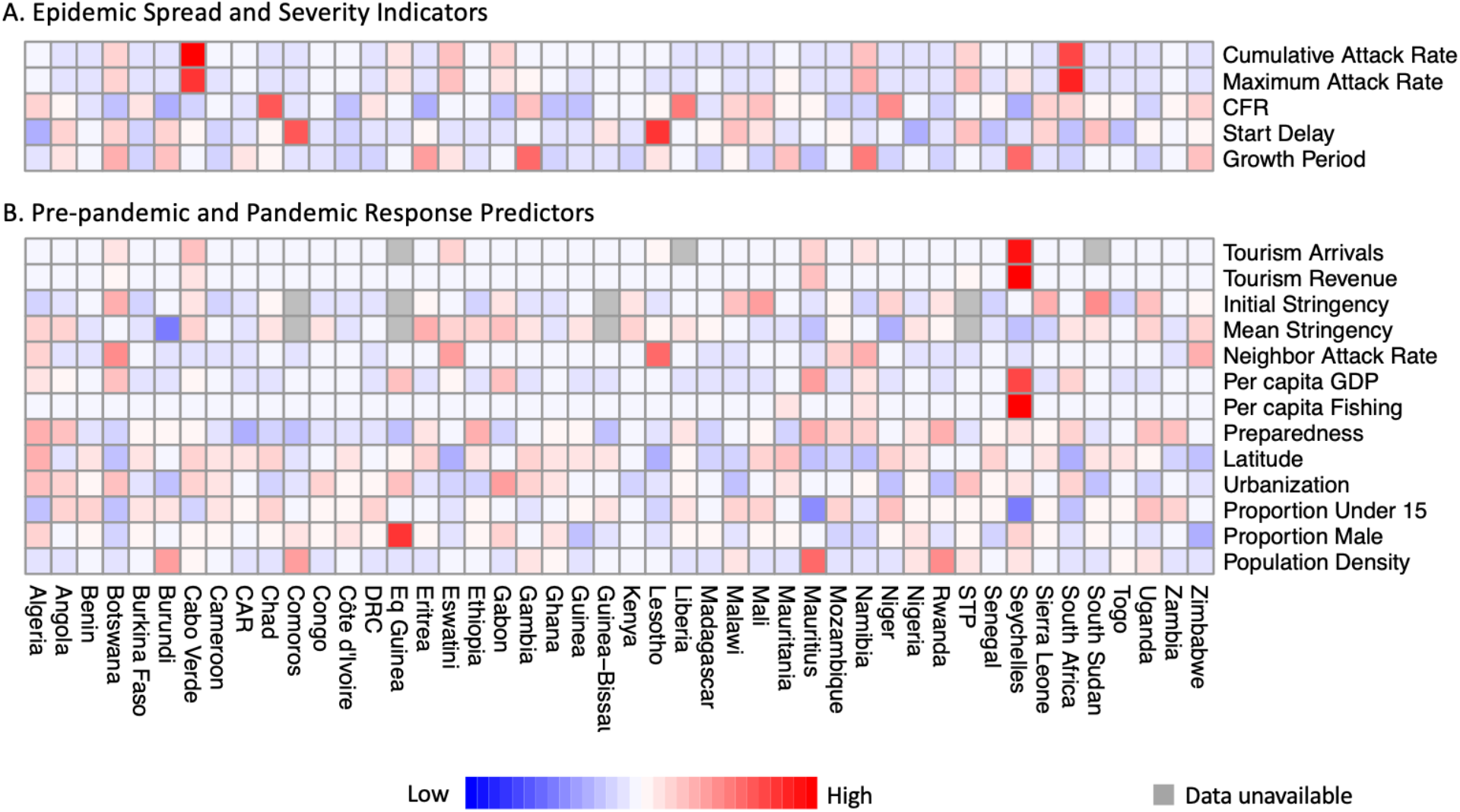
Heatmap showing values for (**A**) the 5 epidemic spread and severity indicators and (**B**) the 13 pre-pandemic and pandemic response explanatory variables from the 46 WHO African countries reporting cases. Blue represents high values, red represents low values, and gray shading corresponds to missing data. Each indicator (in the sets of response and predictor variables) here is scaled by the standard deviation and centered by subtracting the mean before plotting.

### Pre-pandemic and pandemic predictor variables

Variation in the values of pre-pandemic and pandemic response predictor variables for the 46 WHO African region member states is illustrated in Figure 2B. Pairwise correlations between predictor variables showed that wealthier economies had larger tourism industries and older populations, and that urbanization was greater in wealthier countries (Figure S4). All other pairwise correlation coefficients were below 0.6 in absolute magnitude. Following PCA, we reduced the number of informative predictor variables to the first four dimensions (Figure 3A), accounting for 72.2% of the variance of the data. Figure 3A shows the contributions of all principal components dimensions, including PC5, which explained an additional 7.7% of the variance, but had an Eigenvalue of just under 1 and loaded with redundant predictor variables (Figure S5). The geographic distributions of PC1 - PC4 values are illustrated in Figure S2B.

**Figure 3.**
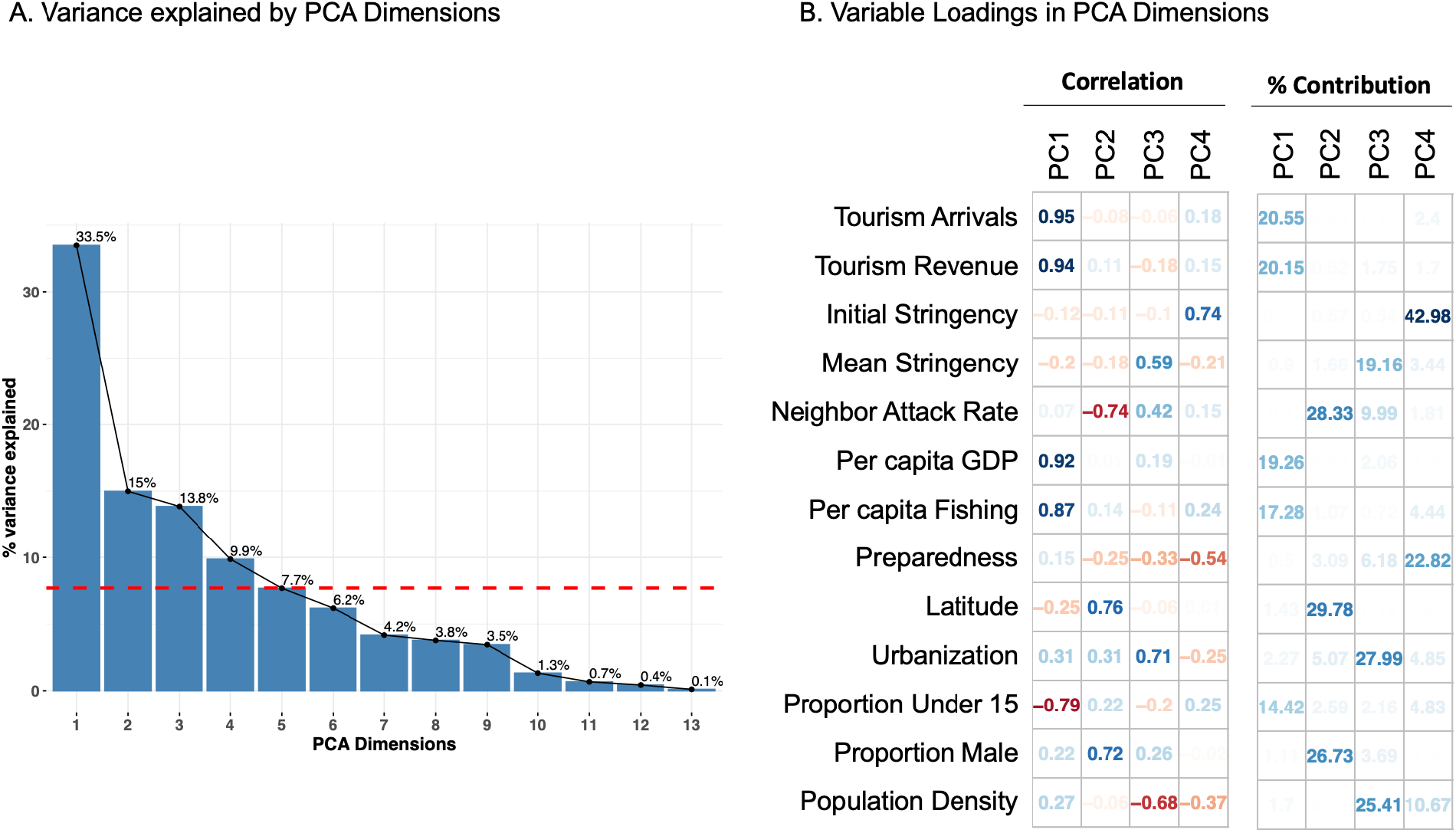
**A:** Scree plot depicting the percentage of variance explained by each PCA dimension. The red line differentiates the four principal components with eigenvalue > 1. **B:** Correlation of each predictor variable with the first four PCA dimensions, and their percent contributions to each of the dimensions. Red refers to negative correlations while blue refers to positive correlations. Darker shades imply stronger correlations and contributions.

The first dimension (PC1) accounted for 33.5% of the explained variance. As illustrated in Figure 3B, PC1 was higher in countries with higher income and visitors from tourism, higher per capita GDP, higher per capita fishing volume, and older population structure (p < 0.001 for all). The second dimension (PC2) accounted for 15.0% of the total variance and was positively correlated with higher latitude and larger proportion of males, and negatively correlated with the cumulative attack rate in neighboring countries (p < 0.001 for all). PC3 accounted for 13.8% and was positively correlated with higher urbanization, lower population density, and higher mean COVID-19 government response stringency (p < 0.001 for all). PC4 accounted for 9.9% of the explained variance, and was positively correlated with higher initial government response stringency and lower pandemic preparedness (p < 0.001 for both).

### Regression analyses

Results of the regression analyses are illustrated in Figure 4 and detailed in Table S2. Both the cumulative attack rate and monthly attack rate were highly positively related to PC3 (higher urbanization, lower overall population density, and higher mean stringency; p < 0.001) and PC1 (high tourism, high per capita GDP, high fishing volume, older population; p < 0.001), and to a lesser extent, negatively related to PC4 (lower preparedness and higher initial stringency; p <0.02; Figure 4). Case fatality ratio was negatively influenced by PC1 (p = 0.021), meaning that CFRs were lower in wealthier member states with large tourism and fishing industries, despite those also being the countries with older populations. This relationship was also evident in the pairwise correlation matrix between response indicators, where attack rates were weakly negatively correlated to CFR (Figure S3). The delay in the detection of the first COVID-19 case relative to its first detection in the region, the *epidemic start delay*, was longer in countries with higher values of PC4 (more stringent COVID-19 control measures and lower preparedness, p < 0.002) and lower values of PC2 (p = 0.008), referring to lower latitude, higher attack rate among neighboring countries, and lower proportion of males in the population. The initial epidemic growth period was more protracted in countries with higher values of PC1 (high tourism, high per capita GDP, high fishing volume per capita, older population; p = 0.023), which corresponded to many of the small island member states (Figure S6). The Benjamini-Hochberg adjustment for multiple tests did not result in any qualitative change in these results (highest unadjusted significant p-value = 0.023 became 0.026 following adjustment).

**Figure 4:**
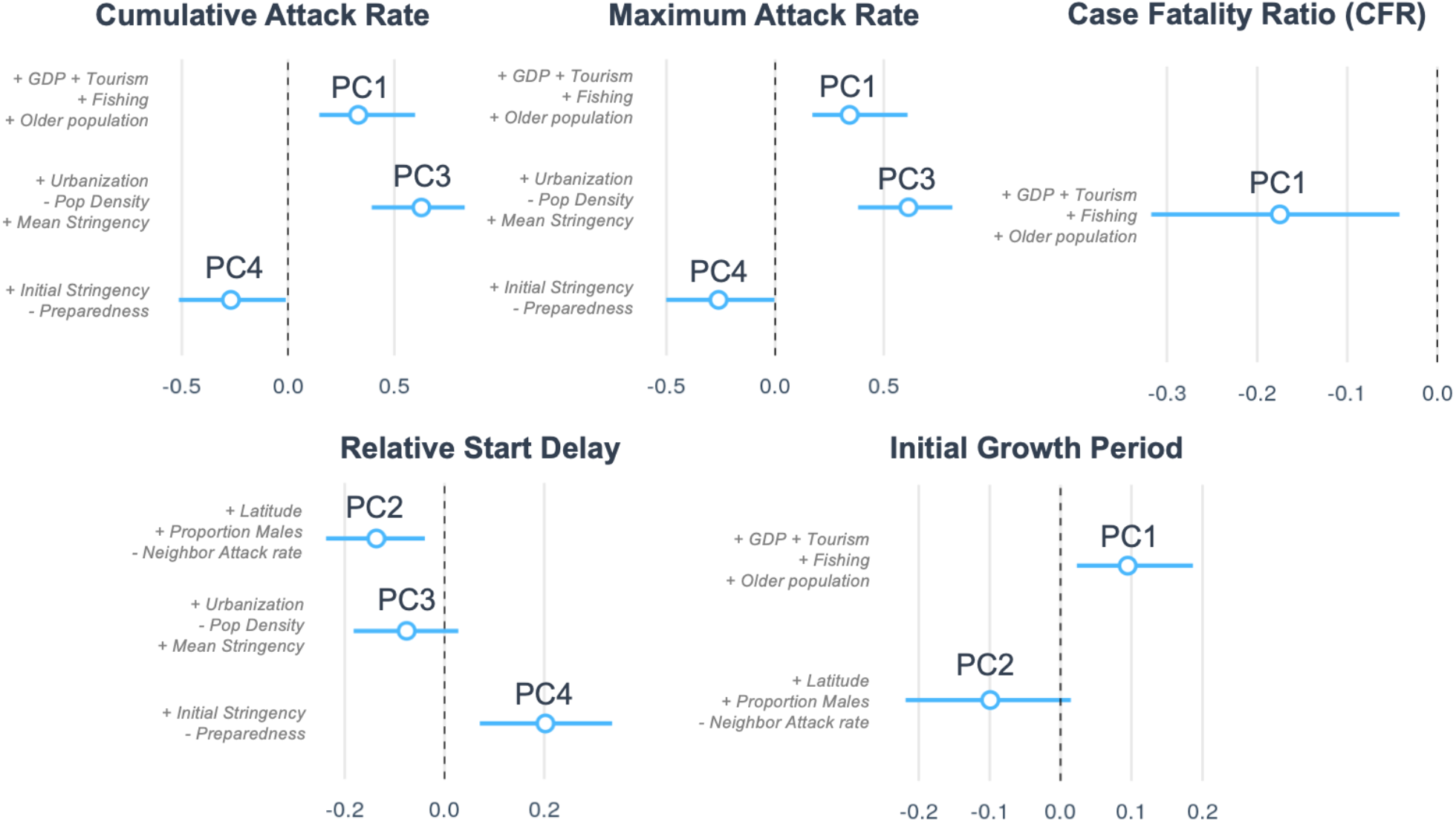
Impact of pre-pandemic and pandemic response predictor variables (summarized as PCA dimensions) on COVID-19 epidemic spread and severity indicators among countries in the WHO African region: The regression coefficients (with 95% confidence intervals) correspond to the best fitting regression model for each epidemic spread and severity indicator. The full regression table is presented in the Supplementary Information Table S2.

### Robustness to data quality

When we re-analyzed the data after removing the countries that had reported fewer than 10 deaths, a proxy we determined was indicative of outliers due to data quality or other factors such as very small population size, we found largely the same results (Appendix S2). While correlations between predictor variables, and thus identity and loadings of principal components, differed, we still recovered very similar results following the regression. Cumulative and peak attack rates were higher, and CFR was lower, in wealthy countries with large tourism industries and older populations. Detection of the first case was more delayed in countries with higher initial stringency scores, and lower values of a composite predictor that loaded heavily with the proportion of males in the population. The only substantive differences between the two analyses were that (1) the initial growth rate (inverse of growth period) was faster in more dense populations, rather than being related to wealth, tourism and age structure; and (2) that fishing volume and mean stringency were not related to any of the response variables in the reduced dataset.

## Discussion

### Major findings and trends

This study considered the geographic, demographic, and socio-economic factors that might have had an impact on the initial (first wave) spread and severity of COVID-19 among 46 of the 47 countries comprising the WHO African region member states. Our primary findings were that 1) wealthier countries with higher per-capita GDP, older populations, and larger fishing and tourism industries reported higher attack rates but experienced slower initial epidemic growth rates and lower case fatality ratios; and 2) countries with lower self-assessment of pre-COVID-19 pandemic preparedness and which imposed more stringent control measures at the start of the epidemic saw a significant delay in the detection of their first COVID-19 case, and reported lower cumulative and peak (mean monthly) attack rates. These findings are consistent with other studies which have documented the association of air travel, connectivity, and tourism on the spread [6,2] and mortality [23] associated with COVID-19 infection for sub-Saharan African countries. This result underscores the importance of the design and early implementation of surveillance and control measures at points of entry, including appropriate screening and isolation policies [11], since these could result in reducing pathogen spread [16,24-26]. Our finding that wealthier countries reported higher (and earlier) case numbers is also consistent with, and partially explained by, heterogeneity in the testing capacity between countries [2], which likely resulted in underreporting or delayed reporting of cases in more resource-limited African countries. When higher attack rates correlate with lower CFR, a trend we observed here, this could be indicative of an impact of capacity on the response in terms of both case detection and clinical management. More directed testing policies in resource-limited settings, such as testing only symptomatic individuals or those in quarantine, may have also contributed to biases complicating the evaluation of actual rates of community spread and severity [27].

It is unclear whether lower numbers of reported COVID-19 cases and deaths in African countries relative to the rest of the world represented lower rates of transmission due to control measures, fewer symptomatic infections due to younger populations, or simply weaker detection capacities than in high-resource settings [28]. It has been well-known from early on that COVID-19 is more severe in older populations [28], driving up the probability of detection. Indeed, African countries were also found to have reported fewer COVID-19 infections among children than adults [29]. However, countries with higher GDP per capita have higher life expectancy at birth [30], suggesting that lower CFRs in better-resourced sub-Saharan African countries could have been attributed to more developed healthcare systems and surveillance capacities [27]. This result presents a paradox, since case fatality rates should be higher in older populations. The explanation we propose is that the paradox itself supports the interpretation that GDP, and therefore surveillance and/or healthcare capacity, is what drove lower CFRs relative to other African countries. In low-resource settings, even if the population is younger, testing capacity is often stretched to the point where it is reserved only for symptomatic or hospitalized cases. Countries with more capacity for contact tracing and systematic testing (eg, in Seychelles) will be significantly less biased towards identifying only the most severe cases, and earlier treatment can lead to better outcomes. That said, high-income countries from other parts of the world, such as the United States, were not successful in reducing mortalities during the first wave of COVID-19, indicating that developed health systems alone were not sufficient to control COVID-19 related deaths [31]. Thus, other aspects responsible for the better performance of relatively wealthy African countries should be explored in future studies.

We found that higher population urbanization was positively associated with higher cumulative and peak attack rates, as well as earlier detection of the first infection. It has been previously recorded that large, global cities reported positive COVID-19 cases at an earlier stage, whether due to their strong connectedness to other large global cities via international travel [32] or due to the concentration of testing resources in capital cities. Additionally, we found that the speed at which the first 50 infections were detected in a country was slower for wealthier countries with larger tourism industries and older populations. This result is counter-intuitive given the likely role of greater detection capacity in these countries, but could indicate that better surveillance may have also led to more effective isolation of early cases. After outliers including the highly wealthy nation of Seychelles were removed from the analysis to test for robustness, we found that this indicator of early epidemic growth rate was no longer influenced by the composite wealth factor (PC1), but instead was classically faster in more dense populations. Dense urban populations are prone to higher rates of crowding and social interactions, which increases the risk for the spread of directly transmitted aerial infectious agents in particular [33-35].

We observed that countries that imposed less stringent control measures at the start of the pandemic were the ones in which COVID-19 outbreak began, or at least was identified, earlier. As expected, early implementation of restriction and control measures probably resulted in delaying the onset of the outbreak. Previous research showed that a common characteristic among countries with delayed onset was the implementation of effective border measures and various preparedness activities at an early stage [10,36,37]. Conversely, countries that had higher preparedness scores were among the earliest to detect their first case. This is best explained by the fact that the preparedness index includes testing capacity, meaning that countries better prepared to control an epidemic are likely to have enhanced testing and surveillance capabilities, and as a result were able to detect and identify cases earlier. Indeed, a study focusing on the 24 sub-Saharan African countries that were COVID-19 free as of March 30, 2020 documented that only 38% of them had COVID-19 testing capacities [10]. Great examples of the positive impact of prior epidemic response capacity-building efforts on the COVID-19 pandemic has come from countries impacted by Ebola virus threats -particularly those touched by the 2014-2016 outbreak in West Africa [38,39]. Although we found that stringent COVID-19 response policies put in place at the start of the pandemic likely helped to control its initial arrival and spread, many of these measures have been associated with other negative consequences [40-43]. Though epidemiologically circular, mean stringency was higher among countries reporting greater attack rates in our analysis. While this association does not imply that on-going stringency measures did not aid in mitigating the size of the pandemic within African countries, as they very likely did, it does show that these measures are neither preventative nor long-term solutions. Therefore, strict government actions should be implemented carefully. Future studies should address the overall impact of such measures by considering more parameters like quality-adjusted life years (QALYs) and effects on mental health [42-45].

#### Strengths and limitations

Our study is one of the most comprehensive studies describing the first wave of COVID-19 pandemic in Africa. Indicators summarizing the arrival of the COVID-19 pandemic in the WHO African region provide a comparative understanding of the burden, severity, spatial trends, and evolution of the pandemic in member states. Our analyses provide evidence for the roles of tourism, age distribution, and GDP in driving COVID-19 attack rates and CFRs. We also established that countries with strict governmental policies experienced later start (or at least detection) dates of the COVID-19 outbreak, and that countries sharing borders with heavily-affected neighbors nevertheless managed to reduce the speed at which the first 50 infections occurred.

However, wide heterogeneity in response capacity across African countries limits our ability to extract general conclusions that accurately reflect the situation on the ground. An important limitation of this study is the lack of standardization in testing [29] and case notification policies between countries. Similar to other regions, African countries experienced under-reporting of confirmed cases and deaths [4,46]. We also note that the crude case fatality ratio (CFR), which uses the cumulative number of deaths reported over the same period as the cumulative number of cases, does not take into account the inherent lag in death and the reporting of deaths; thus, the CFR may be under-estimated, particularly in countries such as Cabo Verde, where the number of cases was still rising steadily at the end of the study period. Additionally, the availability of individual patient data could have been useful, but such studies are limited in the context of Africa and have so far only addressed the impacts of a few individual-level factors on disease frequency and outcomes [47].

Moreover, we relied on non-contextual (average value) data imputation methods where values were missing, as was the case for tourism and stringency variables for some countries. We also note that some indicators were not reported with consistent frequency for all countries, such as population size and density estimates. There are other potential variables not accounted for in our study such as air pollution, climatic variation, economic inequality (GINI index), diet, etc., that could affect the spread and severity of COVID-19 in the region [6].

### Conclusions

Important evidence can be extracted from our results to inform decision-makers on the factors to be considered when designing their plan to effectively and rapidly control a future outbreak in the African context. Relatively wealthy sub-Saharan African countries, which also have large tourism industries, detected higher case numbers early in the pandemic, suggesting a role for both greater testing capacities but also higher exposure via international travel. However, these nations also experienced lower CFRs, potentially due to higher healthcare capacities, allowing them to better manage care of patients and minimize the number of deaths. Countries with weaker control measures faced earlier COVID-19 outbreaks, and those with greater urbanized and dense populations experienced faster increases in the number of cases at the beginning of the outbreak. These findings stress the need to implement appropriate non-pharmaceutical measures at an early stage, with emphasis on densely populated areas, and popular tourist destinations. Where this implementation is challenging, investments in e.g., infrastructure, local production of essential materials (gloves, masks, soap, etc.), training of personnel, and public health education campaigns during non-crisis periods can help improve preparedness. Finally, surveillance and testing capacities remain a key challenge in the region. Robust surveillance and testing capacities are needed to ensure that public health decisions are based on data that depict the epidemiological situation accurately. The quality and timeliness of data are essential to better evaluate and adjust control measures implemented during the course of an outbreak, to help limit the reliance on blanket or prolonged measures that can have harmful social and economic impacts. We thus urge decision-makers to improve these capacities to ensure rapid response to future threats to public health and economic stability.

## Data Availability

COVID-19 data used in this manuscript is available at https://github.com/JyotiDalal93/COVID-19_SSA/blob/main/Data_sub_saharan_Africa.zip. Moreover, data dictionaries, statistical code etc., that support the findings of this study are available upon reasonable request from the corresponding author.

https://github.com/JyotiDalal93/COVID-19_SSA/blob/main/Data_sub_saharan_Africa.zip

## Contributions

AJ conceived the study, with the support of JD, TK, JLA, and DV. AJ, JD, TK, DV designed the study, with the support of JLA, DCPC, and SBM. AJ, JD, TK, DV participated in weekly meetings and exchanged ideas that were helpful for the progress of the draft. JD acquired and cleaned the response variables data with the support of JLA and AJ. TK collected the predictor variable data with the support of JD, AJ, DV, and JLA. AJ performed the statistical analysis with the support of JD, JLA, and DCPC. JLA, DCPC, FCC provided suggestions on statistical analysis. JD, AJ made the plots with the support of JLA, DCPC, and ICR. TK, AJ conducted the literature review, with the support of JD and JLA. AJ, JD, TK interpreted the data, connected it to evidence from literature, and wrote the initial draft with the support of JLA, DCPC, and DV. AJ, JD, TK, DV wrote the initial draft with the support of JLA, DCPC. AJ, JD improved and finalized the draft with the support of JLA, TK, WN, and PA. JLA, OK, FCC, CBH, FM, CC, BS, and BI critically revised the manuscript for important intellectual content and supervised the study. AJ, JD, JLA and TK revised the study following reviewer comments. All authors contributed to final approval of the version to be submitted. The corresponding author attests that all listed authors meet authorship criteria and that no others meeting the criteria have been omitted. AJ, JD, TK, JLA, and OK are the guarantors.

## Data availability statement

COVID-19 data used in this manuscript is available at https://github.com/JyotiDalal93/COVID-19_SSA/blob/main/Data_sub_saharan_Africa.zip.

Moreover, data dictionaries, statistical code etc., that support the findings of this study are available upon reasonable request from the corresponding author.

## Ethical approval

Publicly available COVID-19 data was used in this study, and therefore, no ethical approval was needed.

## Transparency statement

AJ, JD, TK, JLA, and OK affirm that the manuscript is an honest, accurate, and transparent account of the study being reported; that no important aspects of the study have been omitted; and that any discrepancies from the study as originally planned (and, if relevant, registered) have been explained.

## Patient consent for publication

Not required.

