## Supplementary Information for "An in-depth statistical analysis of the COVID-19 pandemic’s initial spread in the WHO African region"

### Table of Contents

Appendix S1: Additional source information for complex predictor variables.

Figure S1: Model Fit Diagnostics.

Figure S2. Geographic distribution of response indicators and composite predictor variables.

Figure S3: Pairwise correlation among response indicators.

Figure S4. Pairwise correlation among predictor variables.

Figure S5: PCA loadings for first 10 dimensions

Figure S6: PCA biplot among islands, inland and coastal member states

Table S1. Data Definitions and Sources.

Table S2: Regression Results

Appendix S2: Robustness Analysis

#### Appendix S1: Additional source information for complex predictor variables.

**Pandemic preparedness index:** We used the Electronic State Parties Self-Assessment Annual Reporting Tool (e-SPAR) to quantify the level of preparedness of each country to manage the pandemic on a national level. The e-SPAR is a digital platform that enables the State Parties to report annually to the World Health Assembly on the state of the implementation of the capacity requirements regulated by the International Health Regulations (IHR). The IHR is a legal instrument of international law that requires the development and maintenance of basic capacities to prevent, monitor, detect and respond to public health risks and emergencies. The e-SPAR measures 13 core capacities, which include a total of 24 indicators. To build our preparedness index we used 9 of the 13 capacities. We excluded C3 (zoonotic events and the human animal interface), C4 (food safety), C12 (chemical events), C13 (radiation emergencies) as they are not strictly related to a global pandemic caused by a virus with aerosol human-to-human transmission once it has already emerged from its natural source. Each capacity was assigned a score defined by the sum of the scores of its indicators. For each country, the preparedness score was defined as the average of these 13 core capacity scores (World Health Organization, 2005).

**Stringency Index**: We used the Oxford Coronavirus Government Response Tracker (OxCGRT) Stringency Index to quantify the strictness of government policies in response to the COVID-19 pandemic for each country. The stringency index is a composite measure based on nine specific indicators, bounded between 0 and 100 (100 = strictest). The indicators used to produce the index were: 1) school closures, 2) workplace closures, 3) cancellation of public events, 4) restrictions on public gatherings, 5) closures of public transport, 6) stay at home requirements, 7) public information campaigns, 8) restrictions on internal movements, and 9) international travel controls, as listed in the official website of the project. We used two versions of the stringency index for each country: 1) the stringency index score measured on the day the first infection was detected in the country (Initial Stringency) and 2) the difference between the mean stringency over the study period and the minimum stringency value achieved over the study period (Mean Stringency). The difference from baseline was used to calculate the mean because many countries had strict policies concerning the border and travel, even a few months before the first local COVID-19 was detected (Hale et al., 2021).

**Cumulative Attack rates among neighboring countries**: For each of the 46 countries included in our study, we summed the total number of cases reported in its bordering countries, as of 29 November 2020, and divided this number by the sum of the population sizes of those countries. We assigned the value 0 for all the island countries since they do not share land borders.

#### Figure S1: Model Fit Diagnostics.

Residual distributions for the best-fit regression model for each response indicator.

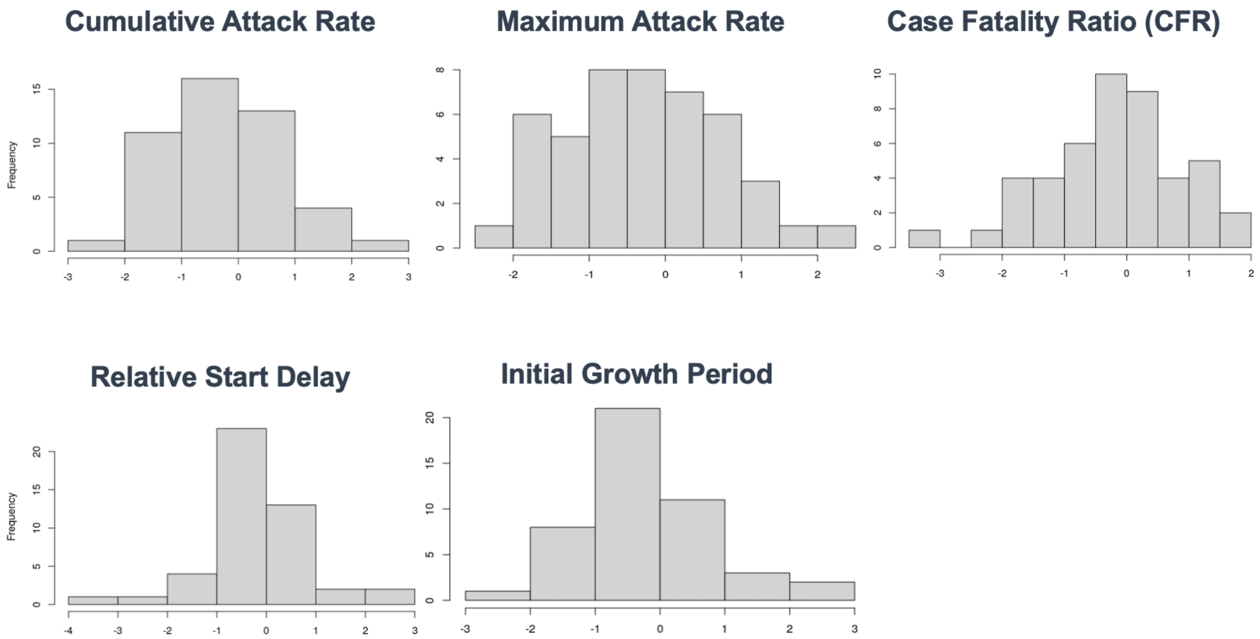

### Figure S2. Geographic distribution of response indicators and composite predictor variables.

The geographic distributions of (A) COVID-19 epidemic spread and severity indicators and (B) principal component (PC) dimensions summarizing 13 pre-pandemic and pandemic response predictor variables for World Health Organization African Region member states. Blue hashing represents countries that were not included in the analysis.

(A) Epidemic spread and severity indicators

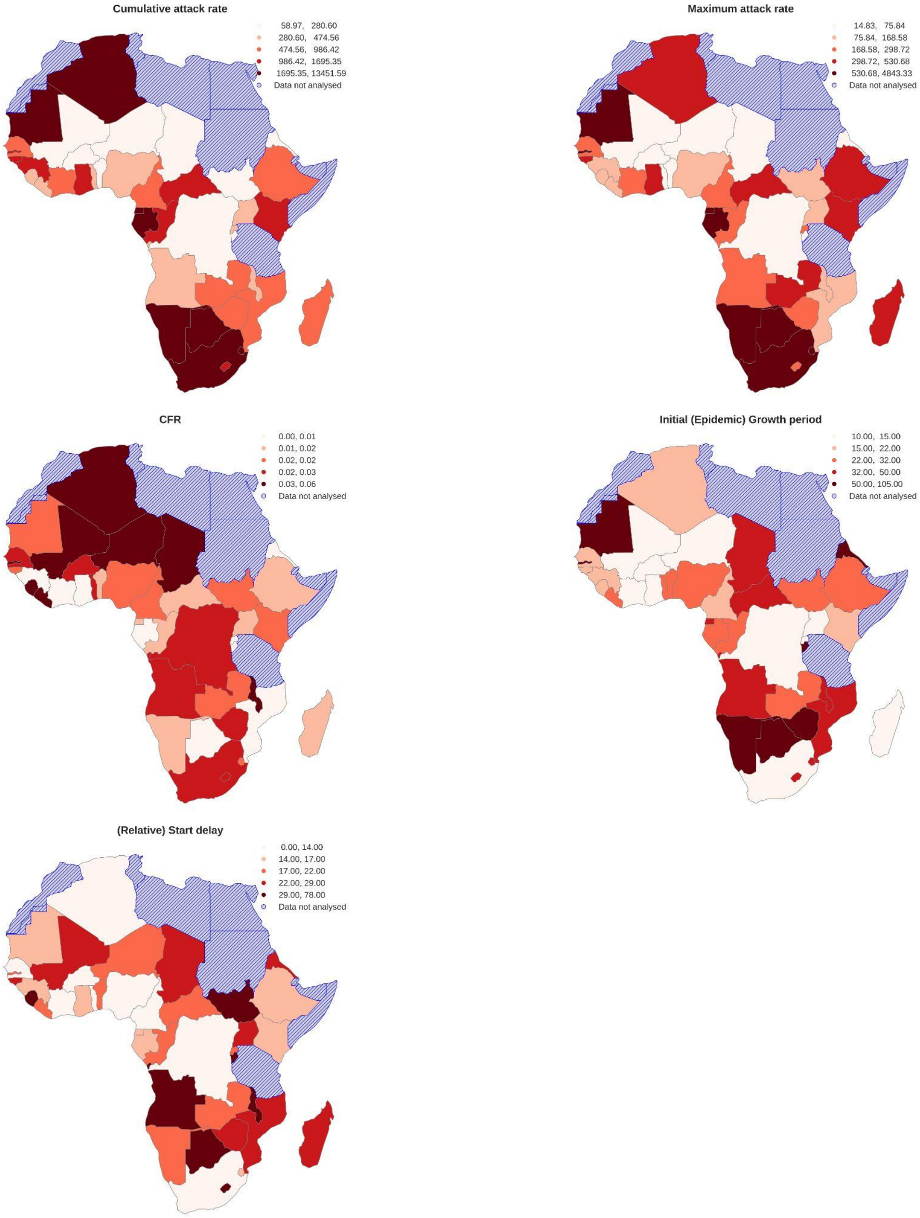

Figure S2 (continued)

(B) Composite predictor variables (PCA dimensions)

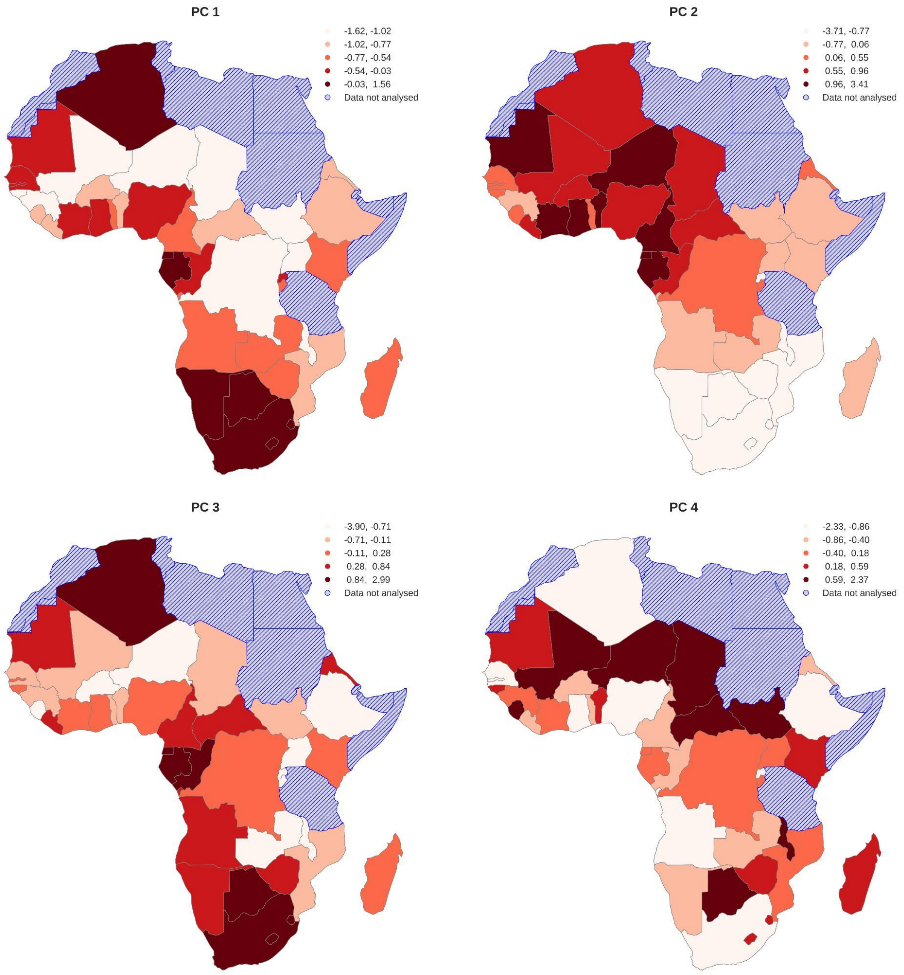

#### Figure S3: Pairwise correlation among response indicators.

Pairwise correlations between epidemic spread and severity (response) indicators. Asterisks (***) indicate $p < 0.001$.

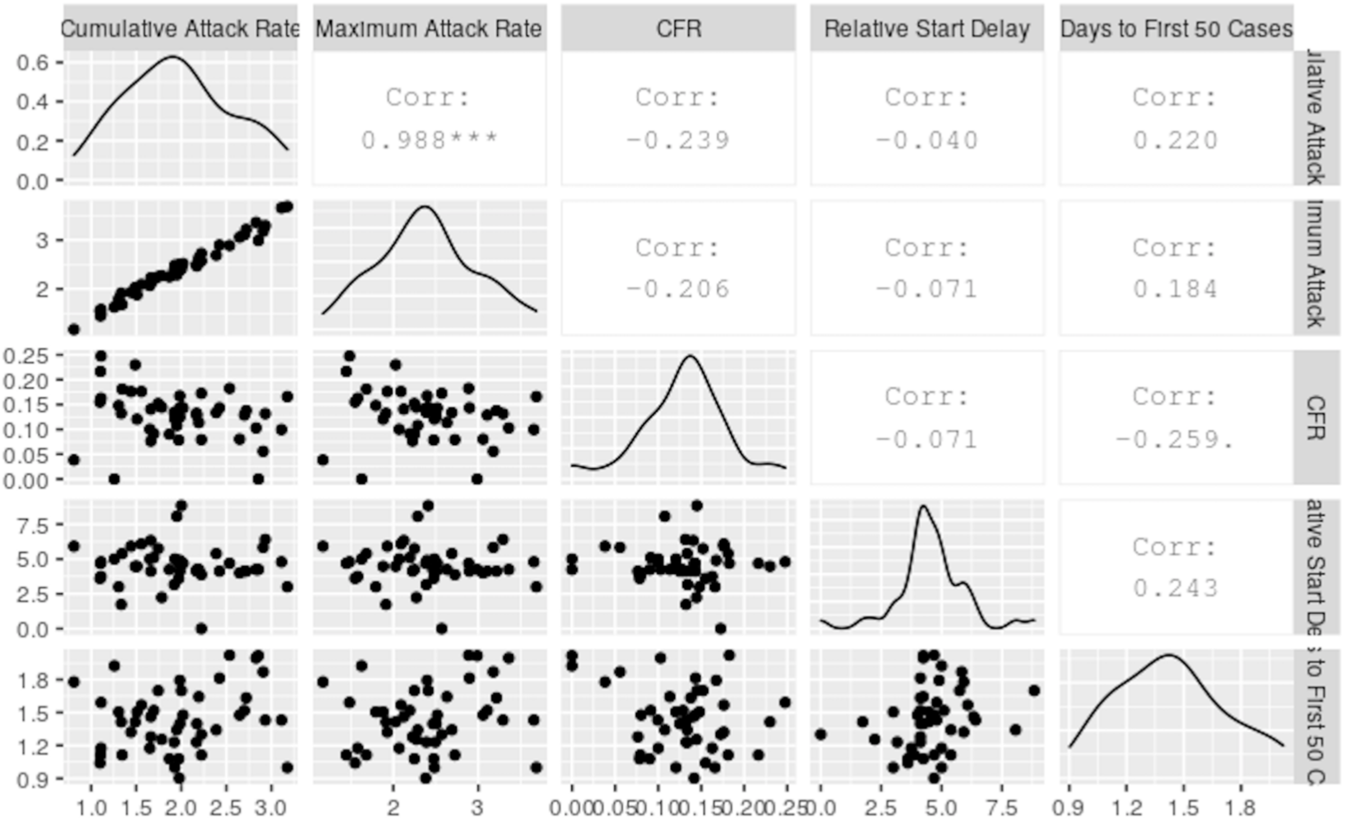

**Figure S4. Pairwise correlation among predictor variables.**

Pairwise correlation matrix depicting the predictor variables included in the PCA, for the full list of 46 countries. The blue color shows a strong positive correlation, whereas red color represents a strong negative correlation. Circles sizes above the diagonal correspond to the numerical values shown below the diagonal, with larger circles for larger correlation coefficients. These numerical values indicate Pearson coefficients, assuming a linear relationship between two variables.

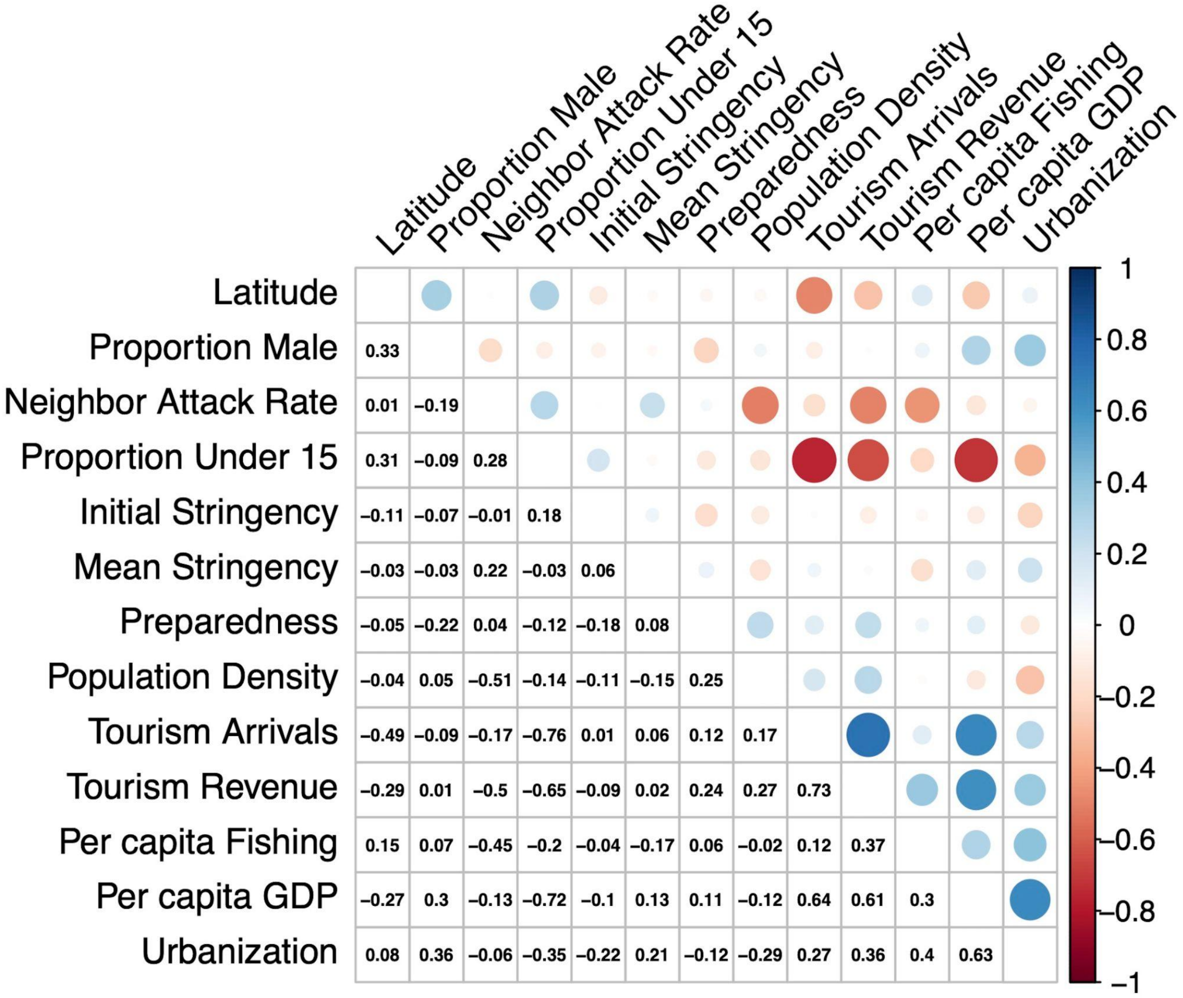

#### Figure S5: PCA loadings for first 10 dimensions

Correlation of each predictor variable with the first 10 PCA dimensions. Red refers to negative correlations while blue refers to positive correlations. Darker shades indicate stronger correlations and contributions.

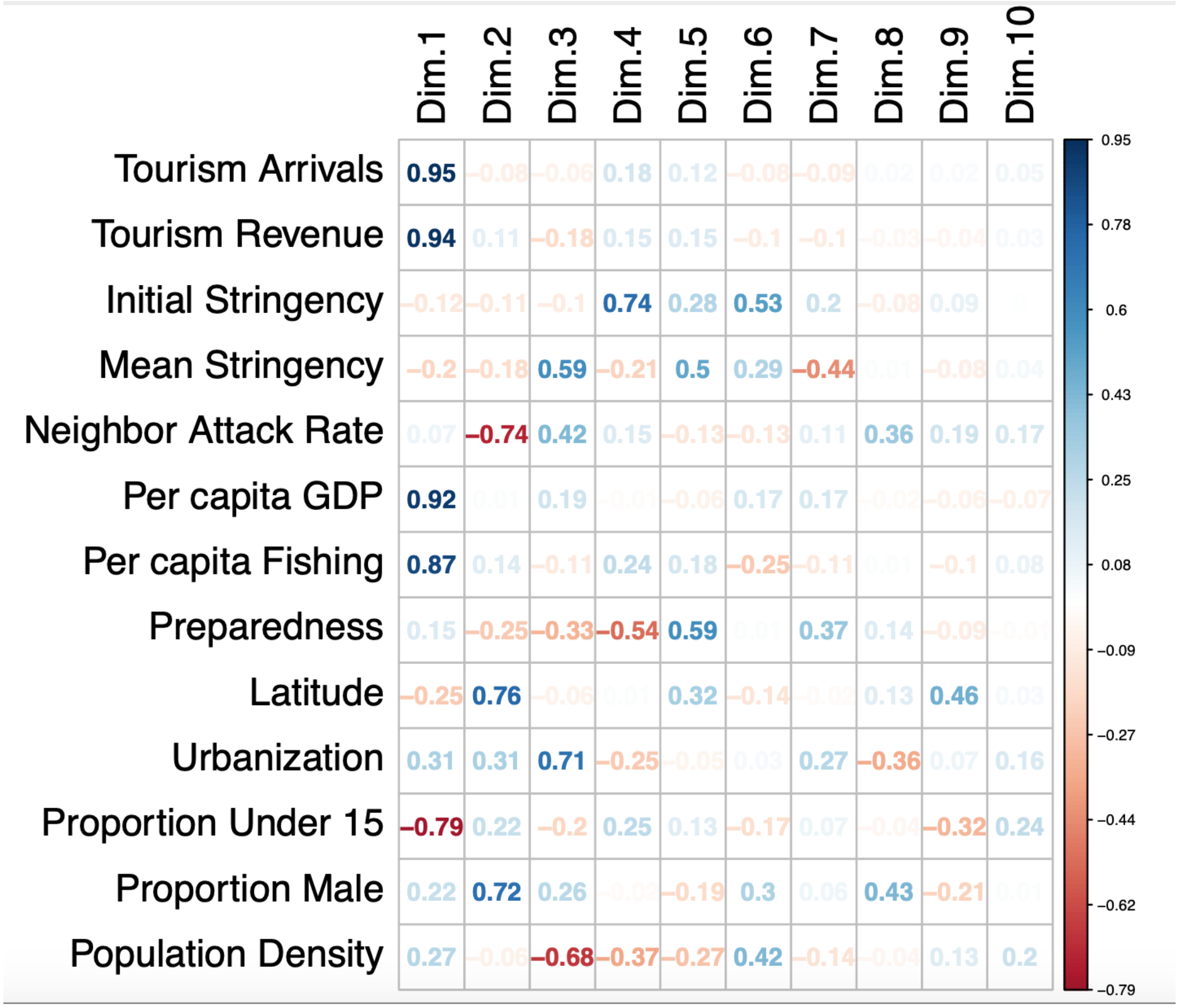

#### Figure S6: PCA biplot among islands, inland and coastal member states

Distribution of inland, island, and coastal WHO African member states across the PC1-PC2 predictor parameter space. Ellipses depict 95% confidence intervals.

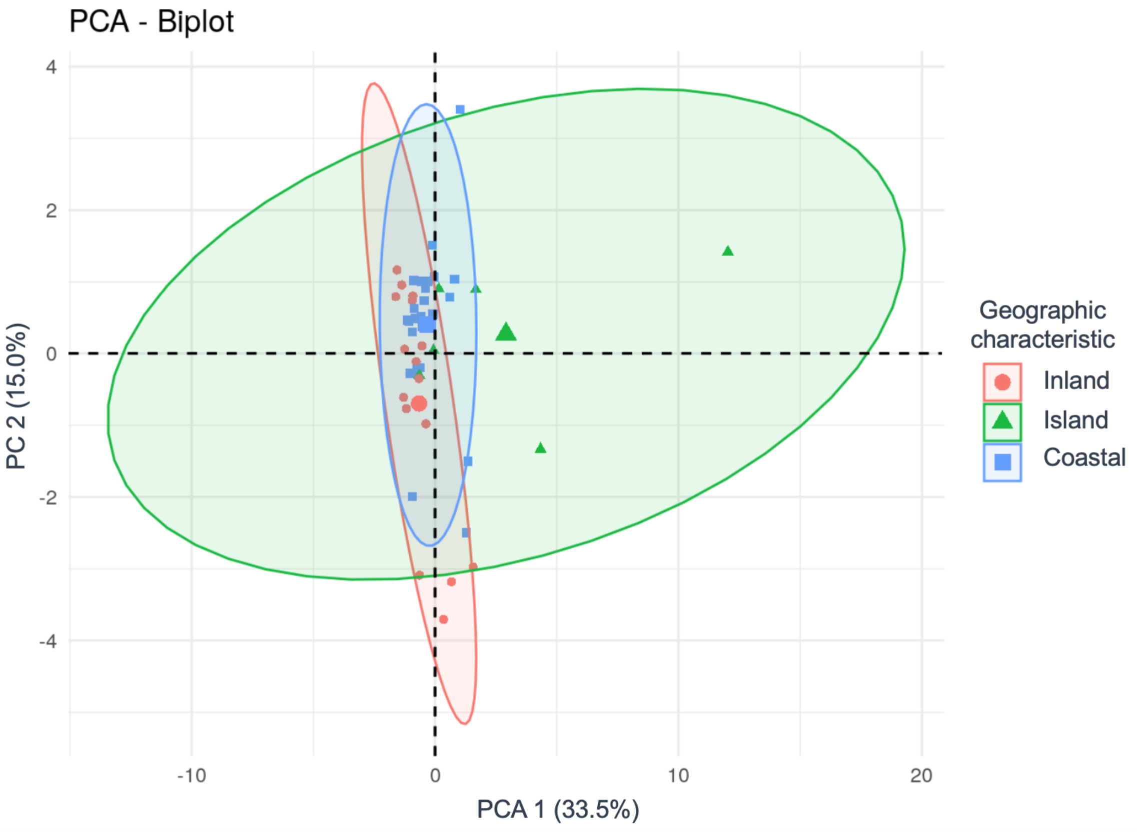

#### Table S1. Data Definitions and Sources.

A list of all variables used in the study and their detailed descriptions and sources.

| Variable | Variable description | Source |
|---|---|---|
| **Epidemic Spread and Severity Indicators** | | |
| Cumulative attack rate | Cumulative attack rate (cumulative number of cases per million inhabitants) | World Health Organization, WHO Coronavirus Dashboard, Accessed 20 Jan 2021. https://covid19.who.int/ |
| Maximum attack rate | Maximum monthly attack rate (new cases per million inhabitants summedcounted over consecutive 4-week intervals) | World Health Organization, WHO Coronavirus Dashboard, Accessed 20 Jan 2021. https://covid19.who.int/ |
| CFR | Crude case fatality ratio (CFR, the ratio of the cumulative number of deaths to the cumulative number of cases, not accounting for lags in death or the reporting of deaths) | World Health Organization, WHO Coronavirus Dashboard, Accessed 20 Jan 2021. https://covid19.who.int/ |
| (Relative) Start delay | The number of days between the date on which first case was detected in each country and the date on which the first case was detected in the region (25 Feb 2020 in Algeria) | World Health Organization, WHO Coronavirus Dashboard, Accessed 20 Jan 2021. https://covid19.who.int/ |
| Initial (Epidemic) Growth period | The length of the initial epidemic growth period, defined as the number of days between the first case and 50th case detected within each country, also expressed as the inverse of the epidemic growth rate | World Health Organization, WHO Coronavirus Dashboard, Accessed 20 Jan 2021. https://covid19.who.int/ |
| **Pre-pandemic and pandemic response (predictor) variables** | | |
| Neighbor attack rate | Combined cumulative COVID-19 attack rate in neighboring countries | World Health Organization, WHO Coronavirus Dashboard, https://covid19.who.int/ ; Our World In Data Coronavirus case database, https://ourworldindata.org/covid-cases Accessed 20 Jan 2021 |

| | | |
|---|---|---|
| Per capita GDP | Per capita Gross Domestic Product (GDP) | The World Bank, https://data.worldbank.org/ |
| Proportion Under 15 | Proportion of of the population under 15 years of age | Indexmundi, https://www.indexmundi.com/ |
| Proportion Male | Proportion of the population that is male | The World Bank, https://data.worldbank.org/ |
| Population density | Population density | The World Bank, https://data.worldbank.org/ |
| Urbanization | Share of population living in urban areas | The World Bank, https://data.worldbank.org/ |
| Latitude | Latitude | https://developers.google.com/public-data/docs/canonical/countries_csv |
| Tourism Revenue | Per capita revenue generated from tourism | The World Bank, https://data.worldbank.org/ |
| Nb tourist arrivals | Per capita number of tourist arrivals | The World Bank, https://data.worldbank.org/ |
| Per capita Fishing | Size of the Fishing industry (Fishing volume per capita) | The World Bank, https://data.worldbank.org/ |
| Initial stringency | COVID-19 government response stringency index score at the time of reporting of the first case [11], | Hale T., Angrist N., Goldszmidt R., et al. "A global panel database of pandemic policies (Oxford COVID-19 Government Response Tracker)." Nature Human Behaviour. 2021;5, 529–538. DOI: 10.1038/s41562-021-01079 |
| Mean stringency | Mean stringency index score calculated as the difference between the mean and minimum stringency index scores over the course of the study period, | Hale T., Angrist N., Goldszmidt R., et al. "A global panel database of pandemic policies (Oxford COVID-19 Government Response Tracker)." Nature |

| | | |
|---|---|---|
| | | Human Behaviour. 2021;5, 529–538. DOI: 10.1038/s41562-021-01079 |
| Pandemic Preparedness | The country's self-assessed level of epidemic and pandemic preparedness | World Health Organization's e-SPAR dashboard: https://extranet.who.int/e-spar |

#### Table S2: Regression Results

Impact of pre-pandemic and pandemic response predictor variables (summarized as PCA dimensions) on COVID-19 epidemic spread and severity indicators among countries in the WHO African region. Results of Best-fit regression models and coefficients for each of the five response indicators. Asterisks describe significance: *** p < 0.001 '**' for < 0.01, '*' for < 0.05, and '.' for <0.1.

| Response Indicator | Best-fit model | Coefficients | | | |
|---|---|---|---|---|---|
| Cumulative Attack Rate | ~ (PC1 + PC3 + PC4) | Intercept | PC1 | PC3 | PC4 |
| | | **7.065** *** | **0.33** *** | **0.628** *** | **-0.27** * |
| | Std. Error | 0.121 | 0.058 | 0.09 | 0.107 |
| | z-score | 58.338 | 5.687 | 6.953 | -2.523 |
| | p-value | < 2e-16 | 1.29e-08 | 3.57e-12 | 0.0116 |
| Maximum Attack Rate (monthly) | ~ (PC1 + PC3 + PC4) | Intercept | PC1 | PC3 | PC4 |
| | | **5.921** *** | **0.344** *** | **0.614** *** | **-0.261** * |
| | Std. Error | 0.125 | 0.06 | 0.093 | 0.11 |
| | z-score | 47.389 | 5.743 | 6.594 | -2.368 |
| | p-value | < 2e-16 | 9.28e-09 | 4.28e-11 | 0.0179 |
| Case Fatality Ratio | ~ PC1 | Intercept | PC1 | | |
| | | **-4.014** *** | **-0.175** * | | |
| | Std. Error | 0.092 | 0.076 | | |
| | z-score | -43.42 | -2.3 | | |
| | p-value | < 2e-16 | 0.021 | | |
| Start Delay (relative to the region) | ~ (PC2 + PC3 + PC4) | Intercept | PC2 | PC3 | PC4 |
| | | **3.084** *** | **-0.137** ** | **-0.076** | **0.203** ** |
| | Std. Error | 0.073 | 0.052 | 0.054 | 0.064 |
| | z-score | 42.247 | -2.647 | -1.398 | 3.162 |
| | p-value | < 2e-16 | 0.008 | 0.162 | 0.002 |
| Initial Epidemic Growth Period | ~ (PC1 + PC2) | Intercept | PC1 | PC2 | |
| | | **3.5** *** | **0.095** * | **-0.099** | |
| | Std. Error | 0.089 | 0.042 | 0.063 | |
| | z-score | 39.341 | 2.271 | -1.569 | |
| | p-value | < 2e-16 | 0.023 | 0.117 | |

#### Appendix S2: Robustness Analysis

Robustness to data quality: Results from reduced (N=42 countries) dataset (excluding countries reporting fewer than 10 cumulative deaths: Burundi, Comoros, Eritrea, and Seychelles). **A:** Scree plot showing variation explained by principal components. **B:** Loadings (correlation and contribution) of predictor variables for informative PC dimensions. **C:** Results of regression analyses testing for the impact of (composite) predictor variables on the spread and severity of the pandemic in each country.

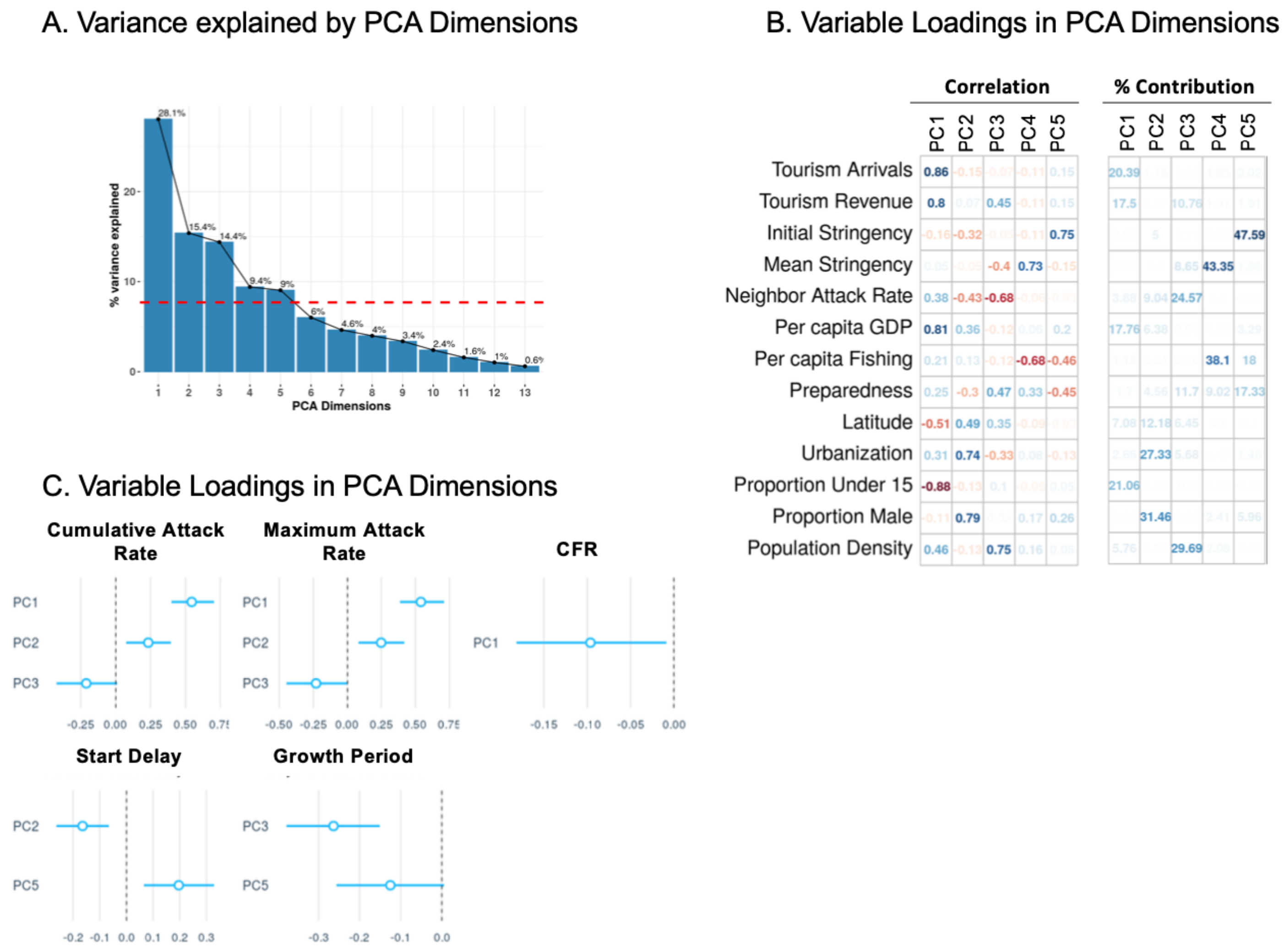


***Principal components analysis:***

The first five principal components had eigenvalues above 1, accounting for 76.4% of the total variance. PC1 (28.1%) captured high per capita wealth, tourism dollars and visitors, and older populations, but was not correlated to fishing volume. The second dimension (PC2), accounting for 15.4% of the total variance, positively varied with the proportion of males ($p < 0.001$) and rate of urbanization ($p < 0.001$). PC3, explaining 14.4% of the total variance, was positively correlated with population density and negatively correlated with attack rates from neighboring countries ($p < 0.001$). PC4, which explained 9.4% of the total variance, correlated positively with mean stringency and negatively with per capita fishing volume ($p < 0.001$ for both). Finally, PC5, explaining 9.0% of the variance, was positively correlated with initial stringency ($p < 0.001$).

***Regression Analysis:***

The cumulative attack rate was positively influenced by PC1 (high tourism, high GDP, and older population; $p < 0.001$) and PC2 (high proportion of males and high urbanization; $p = 0.002$), and negatively related to PC3 (high population density and low attack rates from neighboring countries; $p = 0.008$). The maximum monthly attack rate also showed these same influences. CFR was lower in countries with high PC1 values ($p = 0.045$). Start delay was longer with higher PC5 (high initial stringency; $p = 0.004$) and shorter for PC2 values describing higher urbanization and proportion of males ($p = 0.002$). Finally, the initial growth period was shorter with higher values of PC3 ($p < 0.001$), and trended shorter with higher initial stringency (PC5, $p = 0.08$).
